# Magnitude of wasting and its predictors among under-five children in Bakadawula Ari district, Southern Ethiopia

**DOI:** 10.1101/2023.01.18.23284726

**Authors:** Getamesay Aynalem Tesfaye, Ermias Wabeto Wana, Moges Getie Gebru

## Abstract

**Background:** Globally, wasting threatens the lives of 50 million children under-five. In Ethiopia, wasting is not decreasing at the intended rate, but the reason remains unclear. Moreover, Bakadawula Ari district lacks scientific information regarding wasting among children.

**Objective:** This study was conducted to determine the magnitude of wasting and its predictors among under-five children in the district.

**Methods:** A community-based cross-sectional study was conducted from January to March 2022. A multistage sampling technique was used to select 421 children. The data were entered and analyzed by SPSS 26 (Statistical Package for the Social Sciences version 26). Logistic regression analyses were used and presented with crude odds ratio (COR) and adjusted odds ratio (AOR) with their 95% confidence intervals (CI).

**Results:** The prevalence of wasting among children in the study area was 22.6% (95% CI: 18.5-26.8). Fathers with primary education (AOR= 4.48; 95% CI: 1.93-10.39), households with improper solid waste-disposal (AOR= 2.54; 95% CI: 1.11-5.82), not usually sleeping under insecticide-treated bed net (ITN) (AOR=1.79; 95% CI: 1.01-3.19), unacceptable children dietary diversity score (DDS) (AOR= 2.56; 95% CI: 1.28-5.14) and unacceptable household DDS (AOR= 2.26; 95% CI: 1.02-5.00) were predictors of wasting.

**Conclusions:** The prevalence of wasting among children was critically high. Upgrading educational status of fathers, encouraging safe solid waste disposal, ensuring consistent use of ITN, and improving both children and household DDS should be given a due emphasis to reduce wasting in the study area.

## INTRODUCTION

The term malnutrition refers to all forms of poor nutrition including undernutrition, micronutrient deficiencies, overweight and obesity as well as the resulting diet-related noncommunicable diseases.^1,2^ Wasting, also known as acute malnutrition, occurs in children usually due to insufficient food intake or infectious diseases.^3^ It indicates a recent or current weight loss due to acute under nutrition or illness among children under five, and it is closely associated to increased risk of mortality and functional impairment.^1^

Every country is facing a serious public health challenge from developmental, economic, social, and medical impact of malnutrition.^4^ Globally, around 45% of deaths among children under-five years of age are linked to under nutrition that mostly occurs in low- and middle-income countries.^2^ Children who are undernourished have lowered resistance to infection and are more likely to die from different infectious diseases.^5^ The economic consequences represent losses of 11 percent of gross domestic product every year in Africa and Asia.^4^

In 2020, worldwide 45.4 million (6.7%) under-five children were affected by wasting, two thirds and one quarter of which contributed by Asia and Africa, respectively.^6^ According to Ethiopia Mini Demographic and Health Survey in 2019, 7.2% of children in Ethiopia were wasted.^7^ The survey indicated regional variations, with the highest magnitude (21%) of wasted children living in Somali region and the lowest prevalence (2%) of wasted children residing in Addis Ababa city.

The world’s countries have agreed on targets for nutrition, end all forms of malnutrition by 2030, but, in spite of some progress in recent years, they are off track to reach those targets.^4^ Despite the government of Ethiopia joining the global campaign to end malnutrition by 2030 and decreasing wasting rate lately, the prevalence of wasting was still higher than the national rate in some parts of Ethiopia and remains to be public health problem in the country.^7-9^

Substantial number of studies have identified different factors associated with wasting among children. For instance, a study done in Afghanistan identified education level of household heads, income, age of children, history of children with diarrhea in the last two weeks as determinants of wasting.^10^ A study conducted in India found that children having morbidity were at a higher risk developing wasting,^11^ while another study in Pakistan showed that children whose mothers had no education were more likely to be wasted.^12^ Non-access to health services were associated with wasting in Iran.^13^ Different studies in Ethiopia determined that age of child, fever two weeks before survey, socioeconomic status, family size, and paternal control over resources were among factors associated with wasting.^14-16^

Although the 2019 Ethiopia Mini Demographic and Health Survey provided prevalence of wasting in Southern region of Ethiopia as a whole, it did not indicate the magnitude of wasting specifically among children aged 6–59 months in Bakadawula Ari district, South Omo zone. Moreover, factors associated with wasting were not identified in the national survey.^7^ Various studies in the country reported inconsistent finding on wasting.^8,14,16^ Therefore, this study aimed at determining the prevalence of wasting and its determinants among under five children in the study area.

## MATERIALS AND METHODS

### Study area

The study was conducted in Bakadawula Ari district which is located 745 kilo meter south of Addis Ababa, the capital city of the Ethiopia. The district is found in South Omo Zone which is inhabited by 16 indigenous ethnic groups, and is one of the most common tourist destinations in Ethiopia. In 2021, the district had a total population of 47749 comprised of 7454 children aged 6–59 months. The district has nine kebeles (smallest administrative unit in Ethiopia). There are 2 health centers and 5 private clinics in the district.

### Study design and period

A community-based cross-sectional study was conducted from January 2022 to March 2022.

### Study and source population

The source population for this study was all children aged 6 up to 59 months living in Bakadawula Ari district, whereas the study population was all selected children aged 6 up to 59 months children living in Bakadawula Ari district.

### Eligibility and exclusion criteria

All children aged 6 up to 59 months who were permanent residents in Bakadawula Ari district were eligible. Whereas, children aged 6 up to 59 months who had deformity and critical sickness were excluded from the study.

### Sample size calculation

The sample size for this study was determined using a single population proportion formula:

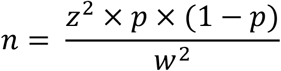

Hence, assuming a 95% confidence level (z=1.96), 5% margin of error(w) and taking 21.2% wasting prevalence(p) from another study in Ethiopia,^9^ the formula yielded sample size(n) of 255. Since the study employed multistage sampling method, a design effect of 1.5 was used and the sample size became 383. After adding 10% for non-response rate, the final sample size was 421.

### Sampling procedure

A multistage sampling method was used, and first three kebeles were selected by simple random sampling technique from all kebeles in the district. Then, the sample size was allocated proportionally to each kebeles based on total number of 6 to 59 months old children. Eventually, using sampling frame prepared by health extension workers, lottery technique was applied to select study participants from each kebele. For those households with more than one eligible child, one of the children was selected randomly.

### Data collection

A validated structured interviewer-administered questionnaire, adapted from available literatures, was used.^17-18^ The questionnaire was composed of questions about socio-demographic factors, health, dietary intakes and environmental factors and it was first prepared in English language and translated into Amharic language then back to English, by two fluent speakers to check the consistency. The data were collected by interviewing the mothers/caretakers, while anthropometric measurements were taken by weighing and measuring height/length of the children. Children were weighed without clothes and shoes using seca 874 (seca, Germany) digital weighing scale. Weight was taken to the nearest 0.1 kilogram. Height/length was measured after taking off shoes and cap via portable wooden height/length measuring board by ensuring the head, shoulders, buttocks, knees and heels of the children touch the measuring board. Length while lying down were measured for children younger than 24 months, whereas, older children’s height was measured while standing. Height/length was measured to the nearest 0.1 centimeter.

### Study variables

#### Dependent variable

Prevalence of wasting among under five children (either wasted or not wasted).

#### Independent variables

Child’s sex, average family size, educational status of father, sex of household head, wealth index, household waste disposal practice, hand washing practice, fever in the last two weeks, diarrhea in the last two weeks, taking the child to health facility during sickness, sleeping under ITN, child’s age at complementary food introduction, child’s meal frequency per day, child’s DDS and household’s DDS.

### Operational definitions

#### Wasting

The percentage of children suffering from moderate or severe wasting (below -2 standard deviation from median weight-for-height (WFH) of reference population).^19^ **Children dietary diversity score:** It is calculated from the number of food items consumed, 24 hours prior to survey date (24 hours recall was used), by the children from the following eight recommended food groups: grain, roots or tubers; vitamin a rich fruits and vegetables; other fruits and vegetables; meat, poultry, fish, sea foods; eggs; legumes and nuts; breast milk; and dairy products. Hence, a child with 5 or more DDS has acceptable DDS.^20^ **Household Dietary diversity score:** It is computed by summing up the number of food items consumed, 24 hours prior to survey date, by the household from the following twelve recommended food groups: cereals; roots and tubers; vegetables; fruits; meat, poultry, offal; eggs; fish and seafood; pulses, legumes, nuts; milk and milk products; oils/ fats; sugar/honey; and miscellaneous. Consequently, a household with 5 or more DDS has acceptable DDS.^20^

### Data quality assurance

All data collectors and supervisors were given two days training on study objective, ethical considerations, ways to approach the study participant and methods of taking anthropometric measurements. Questionnaires were tested prior to actual data collection among children (5% of the sample size) in non-selected kebeles to evaluate the reliability of the tool and procedure. After undergoing the pretest, appropriate change was made on the questionnaire. The weight scale was monitored for its accuracy using an object with known weight. The investigators and the supervisors were checking the completeness of each questionnaire on daily basis. Two data clerks entered the data and consistency was cross checked by comparing the two separately entered data. The data was explored by SPSS v26 and normality of data distribution were checked. Multivariable logistic regression analysis was run to control the confounding factors. Finally, the fitness of the model was tested by Hosmer-Lemeshow’s goodness of fit test.

### Data processing and analysis

The data was entered into and analyzed by SPSS v26 after coding and cleaning. The anthropometric index (WFH) was calculated using ENA for SMART (Emergency Nutrition Assessment for Standardized Monitoring and Assessment of Relief and Transitions) 2020 software. The wealth status of each household was assessed using World Health Organization recommended wealth index construction method by principal component analysis and the wealth status were divided into five quintiles; namely: first (lowest), third (second lowest), third (middle), fourth (second highest), fifth (highest).^21^ Descriptive statistical analysis such as simple frequencies, measures of central tendency and measures of variability were used to describe the characteristics of participants. Then the information was presented using frequencies, summary measures, and tables.

The outcome variable was re-coded to dichotomous outcomes: either they are wasted or not. Those children who are wasted (WFH <-2 standard deviation) were coded as 1, whereas those who are not wasted (WFH >-2 standard deviation) were coded as 0. The independent variables were coded based on previous related studies. All covariates with p value< 0.25 in bivariate logistic regression analysis were considered for further multivariate analysis. Covariates with p-value less than 0.05 during multivariate logistic regression analysis were considered significantly associated with wasting. The logistic regression analysis results were expressed by COR and AOR with their respective 95% CIs.

### Ethical considerations

Ethical approval letter was received from Jinka University Research Ethics Review Committee (reference number: JKU/RCE/ERC/001/14). Confidentiality was assured and mothers/caretakers signed on informed consent form.

## RESULTS

### Socio-demographic characteristics

Four hundred and twenty-one children aged 6 to 59 months were participated. The average age of the children was 22.94 months with standard deviation of 14.28 months. The average age of mothers/caretakers of the children was 27.69 years with standard deviation of 4.83 years. Majority (91.4%) of the mothers/caretakers belong to Ari ethnic group, while 302 (71.7%) of them were Protestant Christianity followers. Regarding educational status, 246 (58.4%) of mothers/caretakers of the children had no formal education (Table 1).

**Table 1:**
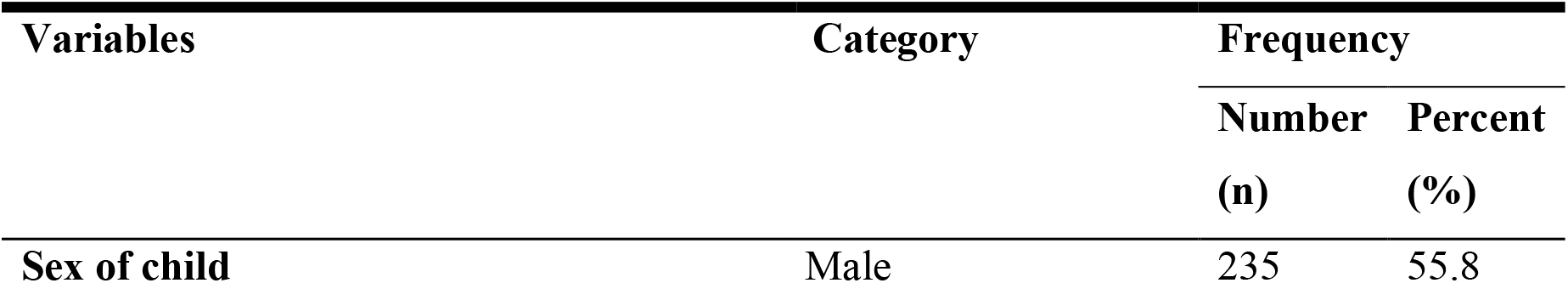
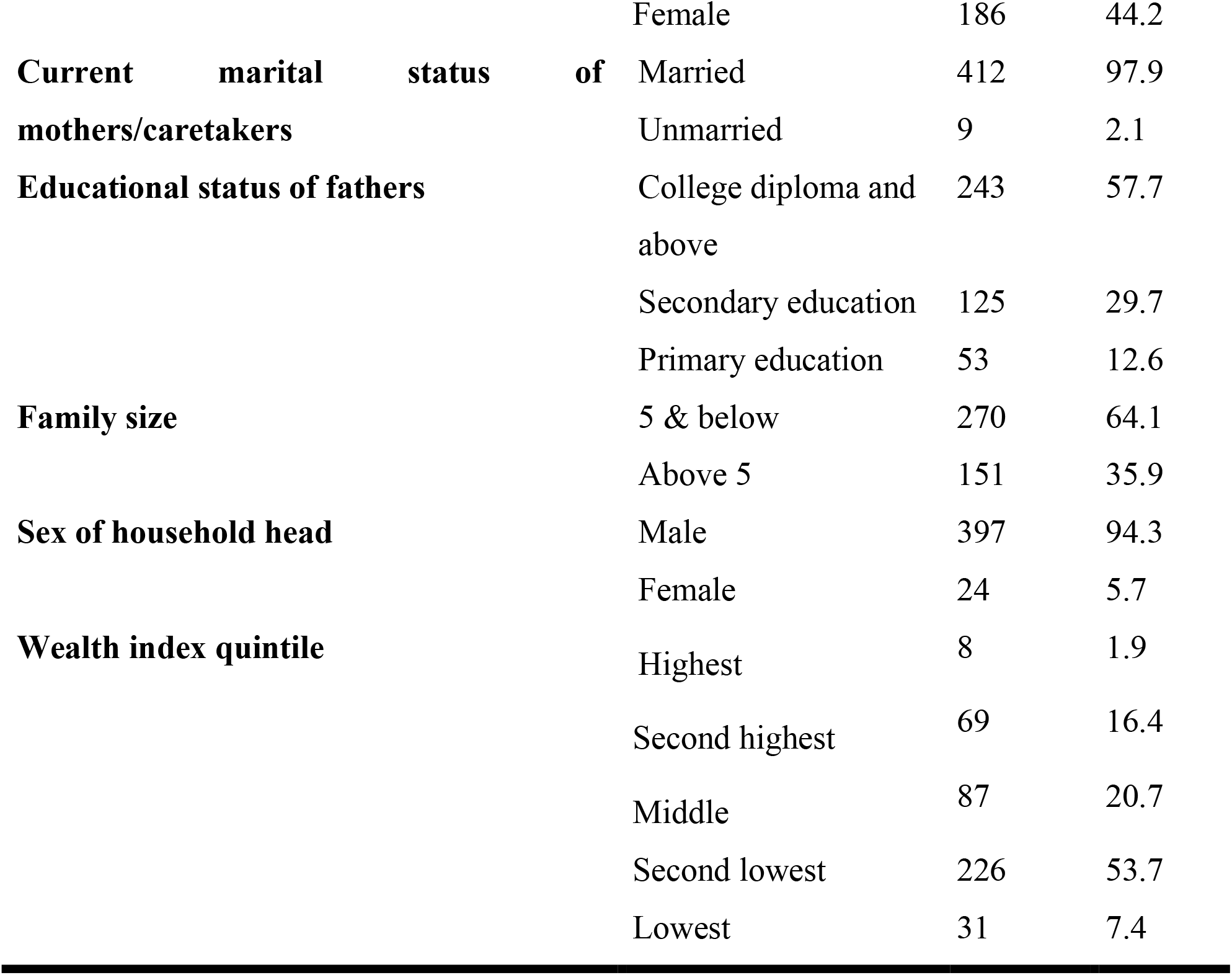
Socio-demographic characteristics of study participants

### Hygiene and health related results

Four fifth of mothers/caretakers of the children in the study were washing hand before feeding a child, however three fourth of them were disposing solid waste improperly. Of all children, almost one third of them had fever and diarrhea two weeks prior to the study. More than half (57.2%) of the children were sleeping under ITN (Table 2).

**Table 2:**
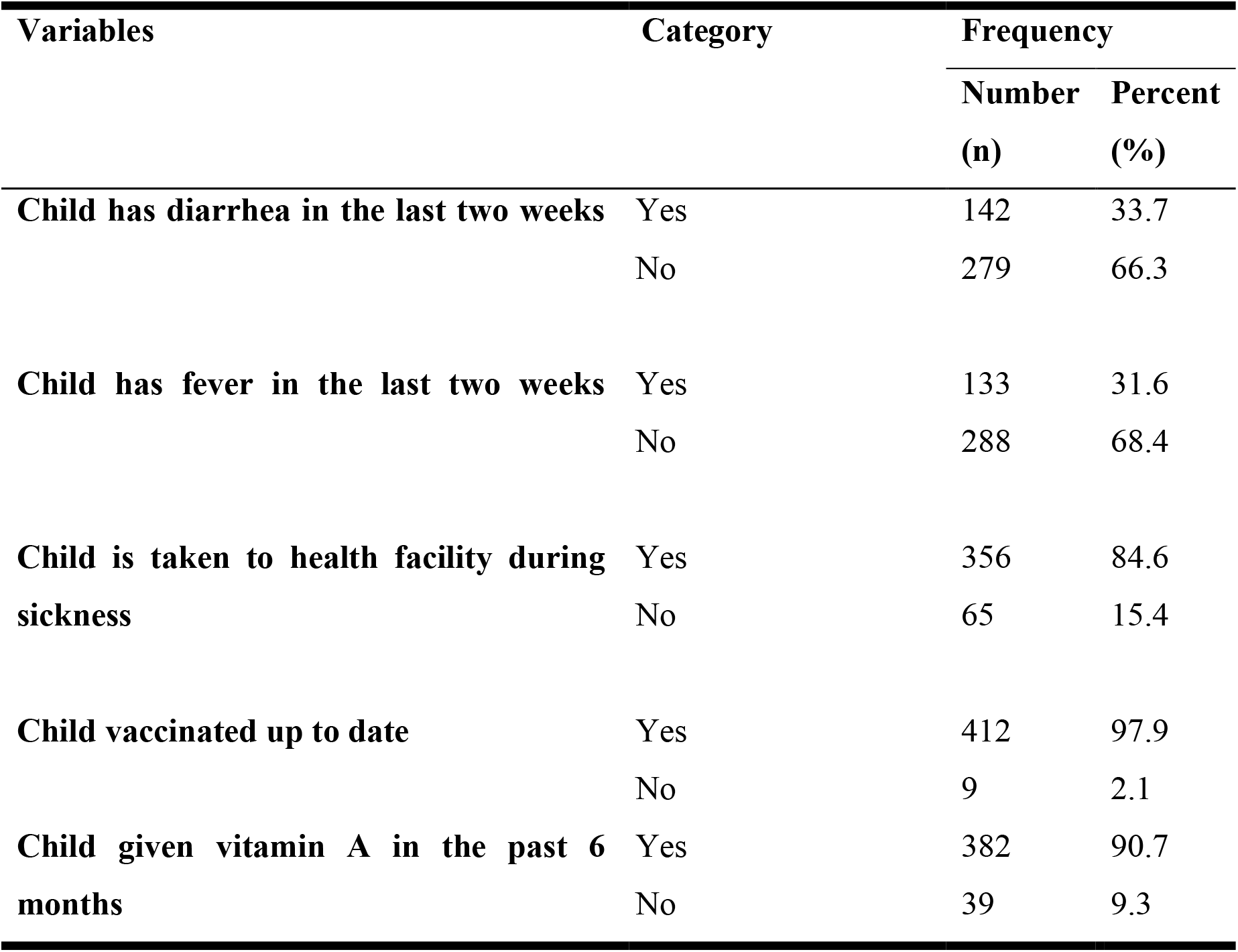
Hygiene and health related attributes of under five children

### Feeding practice and dietary diversity

Most (95.2%) of the children were exclusively breastfed up to 6 months of age, while 373 (88.6%) of them begun complementary feeding at 6 months. Three hundred thirty (78.4%) children had acceptable children DDS. Meanwhile, 359 (85.3%) of households had acceptable household DDS (Table3).

**Table 3:** Feeding practice and dietary diversity of children aged 6-59 months

| Variables | Category | Frequency |  |
| --- | --- | --- | --- |
|  |  | Number<br>(n) | Percent<br>(%) |
| Child begun breastfeeding within 1 hour of birth | Yes | 419 | 99.5 |
|  | No | 2 | 0.5 |
| The number of times the child receives solid, semi-solid, or soft foods on a day before the study | 3 or above | 246 | 58.4 |
|  | <3 | 175 | 41.6 |
| Separately prepare food for child | Yes | 317 | 75.3 |
|  | No | 104 | 24.7 |
| Some foods are restricted (not given) for children in the community | Yes | 2 | 0.5 |
|  | No | 419 | 99.5 |
| Acceptable children dietary diversity score | Yes | 330 | 78.4 |
|  | No | 91 | 21.6 |
| Acceptable household dietary diversity score | Yes | 359 | 85.3 |
|  | No | 62 | 14.7 |

### Magnitude of wasting among the children

The magnitude of wasting (including moderate and severe wasting) among all children was 22.6% (95% CI: 18.5-26.8). The overall magnitude severe wasting was 14.8%, whereas prevalence of moderate wasting was 7.8%. Girls (11.9%) suffered slightly more from wasting than boys (10.8%). The burden of wasting among children aged 6-23 months (26.4%) was more than the rate among children aged 24-59 months (18.7%).

### Factors associated with wasting

The bivariate logistic regression showed that the following fifteen variables were associated, at p-value less than 0.25, with wasting among the children: sex of a child, educational status of fathers, family size, sex of household head, wealth index, solid waste disposal practice, hand washing, child’s diarrheal history, child’s fever history, taking child to health facility during sickness, use of ITN, beginning of complementary feeding at 6 months, the number of times a child receive complementary foods a day before the study, acceptable children DDS, and acceptable household DDS. All of the aforementioned variables did not show any multicollinearity and were candidate for multivariate analysis. Upon multivariate logistic regression, the following five covariates were found to be risk factors of wasting: primary educational status of fathers, improper solid waste disposal, not usually using ITN, unacceptable children DDS and unacceptable household DDS (Table 4).

**Table 4:**
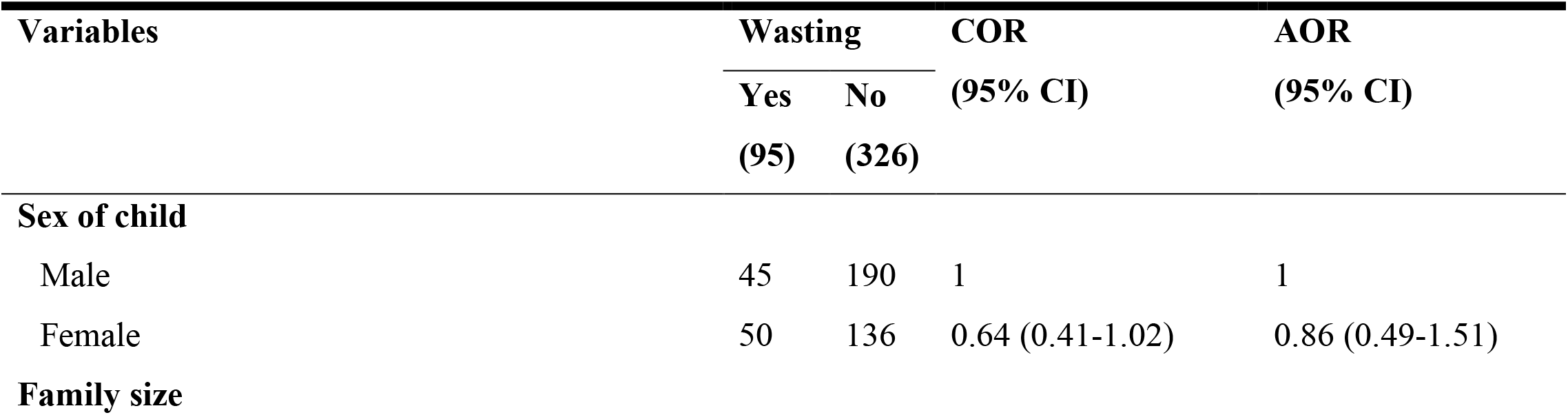
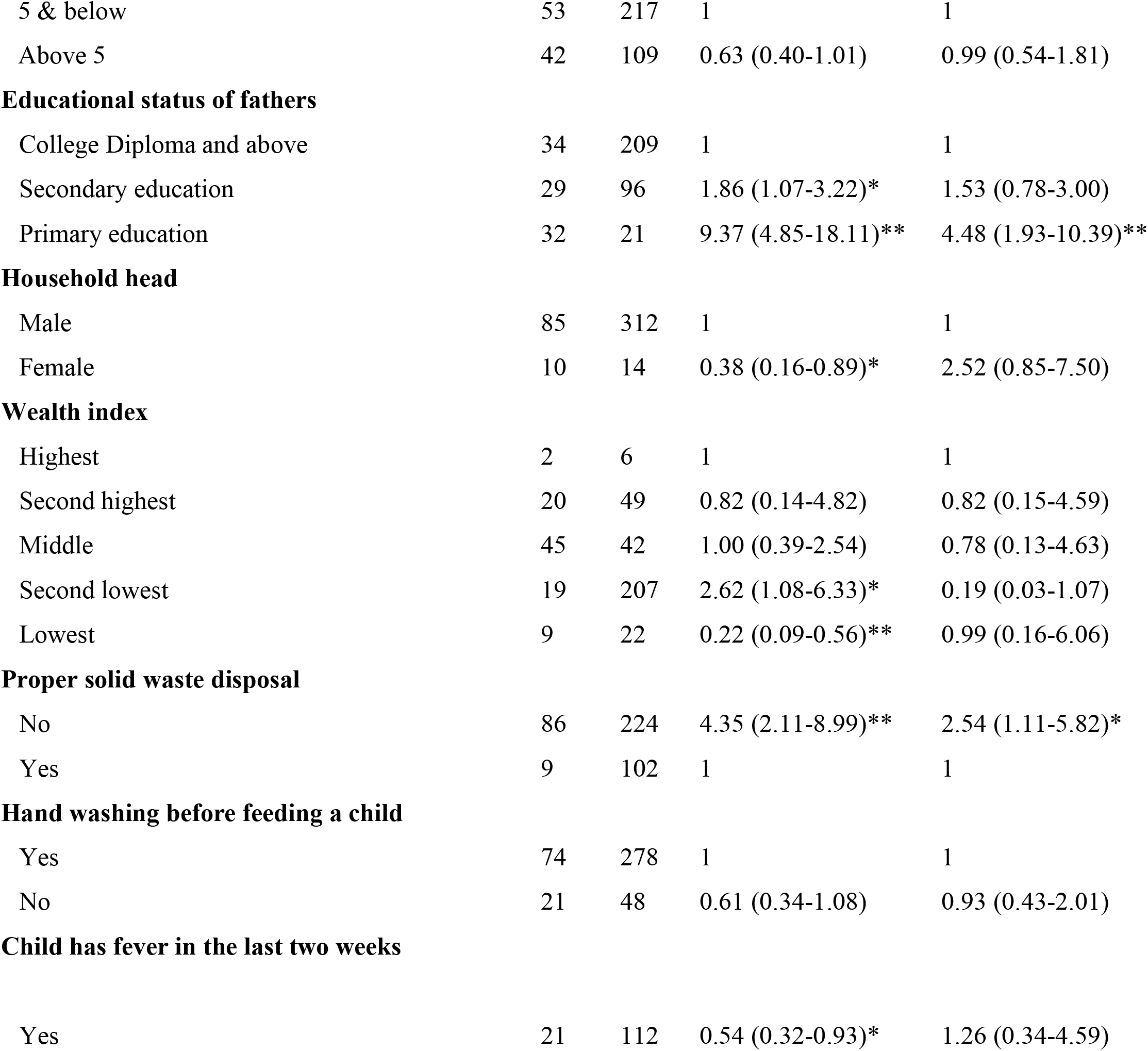
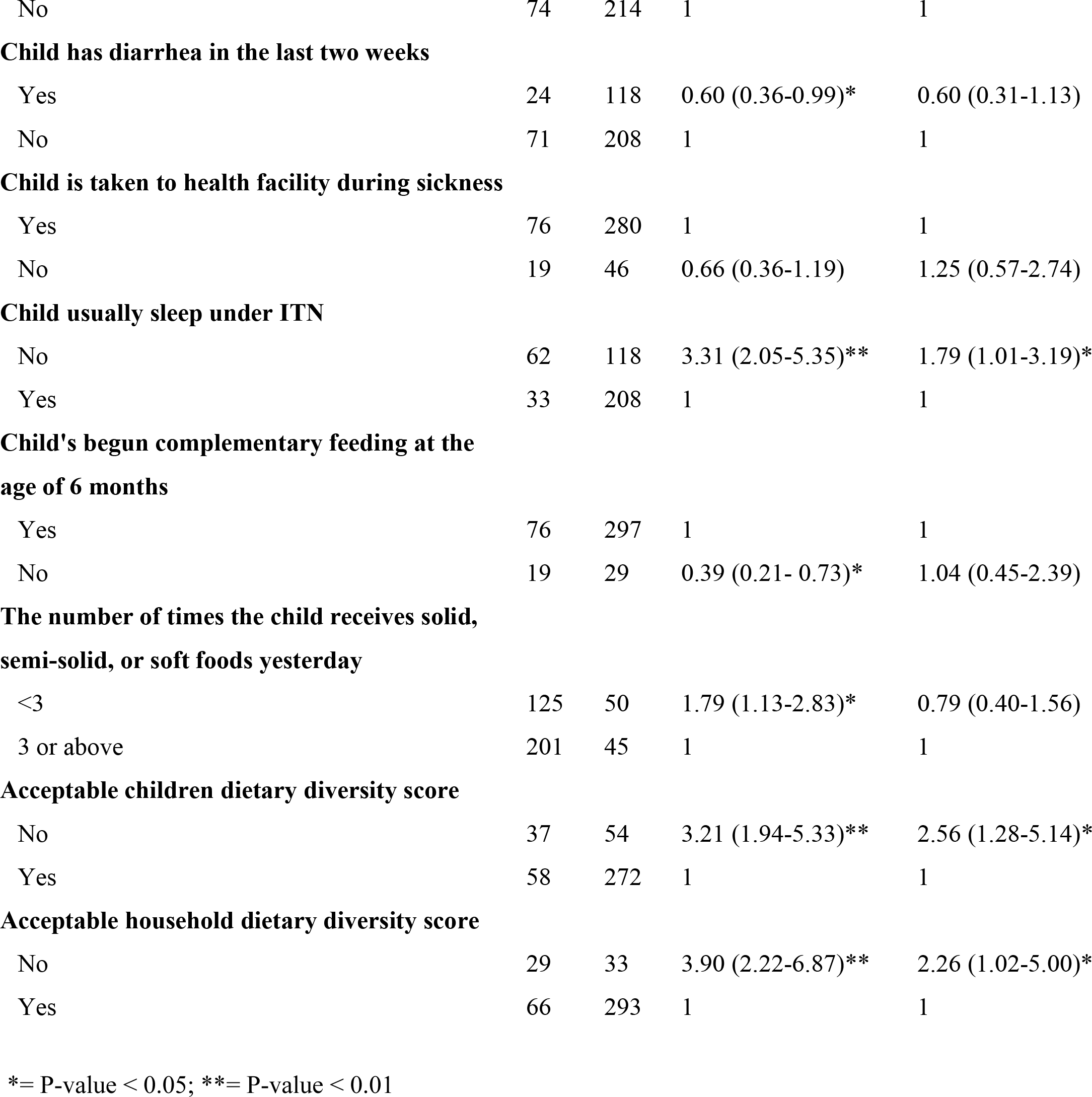
Predictors of wasting among children aged from 6 up to 59 months

Children born to father who had educational status at most primary were 4.48 times (AOR= 4.48; 95% CI: 1.93-10.39) more likely to have wasting compared to those children whose fathers had educational status of college diploma and above. In contrast to under five child whose household dispose solid waste properly, a counterpart had 2.54 times (AOR= 2.54; 95% CI: 1.11-5.82) increased odds of wasting. A child who had not been sleeping under ITN was 1.79 times (AOR=1.79; 95% CI: 1.01-3.19) tends to be wasted than a child who had been sleeping under mosquito bed net. The odds of being wasted among children with no acceptable children DDS were 2.56 times (AOR= 2.56; 95% CI: 1.28-5.14) higher than children with acceptable children DDS. Children with unacceptable household DDS were 2.26 times (AOR= 2.26; 95% CI: 1.02-5.00) more likely to have wasting than children with acceptable household DDS.

## DISCUSSION

The prevalence of wasting among children aged 6–59 months in Bakadawula Ari district was 22.6% (95% CI: 18.5-26.8), which was critically high. It was associated with paternal education, household solid waste disposal practice, children utilization of ITN, child’s DDS and household DDS.

Due to social desirability and recall bias in participants response to the food consumption questions, the study has shortcomings. However, the study was good in studying the magnitude and associated factors wasting simultaneously.

Although the finding was critically high,^3^ it was in line with studies done in Gursum which was 21.2%,^9^ and in Tierkedi South Sudanese refugee camp 23.2%.^22^ However, the current finding was much higher than a study conducted in Addis Ababa 4.7%.^23^ The possible reasons for the discrepancy could be the setting difference where caretakers of children live in; Caretakers in the Addis Ababa were closer to nutrition related information, child care during illness and easily access of market and economically better off than the current study participants. The recent Ethiopian Mini health and demographic survey revealed that seven percent of under-five children were wasted, but the current finding was three-fold higher.^7^ This difference may be due to the national survey reported national average where higher proportion in some areas masked by very smaller proportions in some other parts of the country. Elsewhere, studies done in Kenya and in Nigeria also revealed lower magnitude of wasting than this study, which were 7.1% and 8.8%, respectively.^24-25^ The gaps may be due to recent lack of rain and drought in the current study area which could play pivotal role indirectly to this critically high prevalence of wasting by decreasing production of cereals and crops as well as affecting household livelihood security. Another reason for the discrepancy might be difference in the study settings and time.

As to the current study, children from mothers/care givers whose partners had no to primary education had higher odds to be wasted as to compared to children from mothers/care givers whose partner had college diploma and above educational status. This association was supported by studies performed in Ethiopia,^13^ Sudan,^26^ and Bangladesh,^27^ which depicted that lower level of educational status of caregiver’s partner or children’s father was risk factor for wasting among children.

This study also showed that children whose household dispose solid waste improperly were more likely to suffer from wasting than children whose household dispose solid waste properly. Poor hygiene creates room for increment in diarrheal disease occurrence among under-five children by favoring houseflies and other disease-causing vectors that could lead to infection and then wasting.^28-29^

Children who were not usually sleeping under ITN were more prone to be wasted than children who were usually sleeping under ITN. Different studies revealed that prevention of malarial infection among children by using ITN is crucial for minimizing under five malnutrition and mortality.^30-32^

Compared to children with acceptable children DDS, the odds of being wasted among children with unacceptable children DDS were higher more than double. This association was in the same direction with a study conducted in Karat town,^33^ in Kalafo district,^34^ and in Ghana,^35^ which reported that unacceptable children DDS was risk factor for wasting. Moreover, another study done in Tanzania revealed that consumption of a diverse diet was significantly associated with a reduction of wasting in children.^36^.

This study, unlike most studies on wasting, were able to find that household dietary diversity is associated with wasting among under-five children. Hence, children with unacceptable household DDS were more likely to be wasted than those with acceptable household DDS. This is perhaps due to children with a better household dietary diversity, could have a higher probability of getting sufficient balanced dietary intakes.

Therefore, policy makers and other concerned bodies ought to improve both children and household DDS, and endorse proper solid waste disposal. It should be striven to make sure that children sleep under ITNs via making ITNs accessible and affordable, and other strategies. In the future, researchers should better employ case control study design to establish cause effect relationship between wasting and its determinants.

## Data Availability

The data for this research article is available in the Mendeley repository, DOI:10.17632/ myzxkbwtr5.1

https://data.mendeley.com/datasets/myzxkbwtr5/1

## ACKNOWLEDGEMENTS

The authors would like to thank Jinka University for allowing us to carry out this work and material support for data collection. Jinka hospital and Bakadawula Ari district office also deserve credit for their material support. We extend our sincerely gratitude to data collectors, supervisors and study participants.

## AUTHOR CONTRIBUTIONS

GAT and EWW participated in conception and design of the study, supervision of data collection and management, data analysis, interpretation of the findings, and writing the final report. MGG, AKA, and AAM involved in data management and analysis, interpretation of the findings, and writing the final report. All authors read and approved the final manuscript.

